# A Multi-Country Qualitative Evaluation of Rapid Mortality Surveillance During the COVID-19 Pandemic

**DOI:** 10.1101/2025.09.11.25335597

**Authors:** Carlie Congdon, Farnaz Malik, Ruxana Jina, Diana Kumar, Philip W Setel

**Affiliations:** Vital Strategies, 100 Broadway, 4^th^ Floor, New York, NY 10006 USA

## Abstract

**Background:** Low- and middle-income countries without well-functioning civil registration and vital statistics (CRVS) systems struggle to obtain accurate and complete counts of total (i.e., all-cause) mortality, especially during a pandemic. The aim of this evaluation was to assess whether rapid mortality surveillance (RMS) provided timely and useful counts of deaths to inform pandemic response, despite competing demands on government and public health responders, during the COVID-19 pandemic. This is the first published evaluation of efforts to test the feasibility, utility, and impact of efforts to improve or establish mortality surveillance.

**Methods:** We supported 13 low- and middle-income income countries to implement RMS during the first wave of the COVID-19 pandemic. From August to October 2021, we conducted a qualitative assessment of each country’s progress.

**Results:** Analysis of in-depth interviews with 16 respondents in 13 countries surfaced common themes related to facilitators of RMS implementation. These include 1) government ownership and buy-in; 2) data collection and digitization; 3) interagency collaboration and data sharing; 4) data analysis and interpretation; and 5) data use for decision-making.

**Conclusion:** Robust and digitalized CRVS systems can serve the rapid mortality surveillance function well. For locations where digitization and connectivity of systems are still improving, more feasible and fit-for-purpose approaches are needed. These findings should inform the development of CRVS mortality surveillance functions in low- and middle-income countries.

**Thumbnail Sketch:** WHAT IS ALREADY KNOWN ON THIS TOPIC
Accurate and timely measurements of total and excess mortality are essential to quantify the human toll of a pandemic and inform a country’s response—and are needed quickly and at a time when many other demands compete for resources. In 2020 and 2021, when the activities reported here were implemented, the availability and capacity of diagnostic testing for COVID-19 were limited in most low- and middle-income countries. This complicated the consistent diagnosis of confirmed and suspected COVID-19 cases, leaving few options for tracking the impact of the pandemic. Therefore, we partnered with governments to increase the use of total mortality measurement and comparison to historical averages to assess the spread of the epidemic and the impact on population health.

WHAT THIS STUDY ADDS
Despite the value of rapid mortality surveillance (RMS) to pandemic preparedness and response, there are no other reports that deal primarily with the operational and contextual success factors and constraints that affect its implementation in a pandemic context. Here we identify and summarize local views on barriers and facilitators to establishing RMS during the COVID-19 pandemic in 13 low- and middle-income countries. Results from the evaluation of country processes found that implementation challenges were sometimes severe in the pandemic context, making government buy-in and perseverance even more essential to success. Our evaluation provided evidence of the value of measuring total and excess mortality when systems were able to be operationalized. The insights these data provided were leveraged for better response: improved targeting of limited vaccine supply, action plans that aligned with actual pandemic conditions, and evidence-informed informed risk ratings.

HOW THIS STUDY MIGHT AFFECT RESEARCH, PRACTICE OR POLICY
Investments made today in the active follow-up, enumeration and registration of incident deaths and the digitalization of civil registration and vital statistics systems will bolster health security in countries currently lacking the capacity to generate timely all-cause mortality data. Data on total and excess mortality suited for pandemic preparedness and response should be timely and complete (or at least population-representative) to enable public health officials, government leaders, and policymakers to make data-informed decisions about prevention and protection measures during a pandemic. In the interim, even timely facility-based mortality data can provide some view on a pandemic’s trajectory, if not its population impact.

## Introduction

After the World Health Organization declared COVID-19 a global pandemic on March 11, 2020, those involved in pandemic response needed timely measurement of all-cause mortality (i.e., a count of all incident deaths, regardless of cause) as a broad measure of pandemic impact and trajectory [1, 2]. Many countries used diagnostic testing, case reports, and counts of deaths among diagnosed cases, but these measures were not universally available, nor did they provide a full picture.

Many low- and middle-income countries had limited diagnostic testing early in the pandemic. When coupled with challenges consistently identifying confirmed and suspected cases, this left few options aside from attempting to measure total and excess mortality to assess the pandemic’s impact. “Total mortality” encompasses all deaths, from all causes, in a specified period (by age, sex, place of usual residence, and place of death), including deaths directly and indirectly related to the pandemic and unrelated deaths. For example, it includes deaths potentially attributable to pandemic-related disruptions in healthcare or behavior changes. Early in the pandemic, it became apparent that counting total mortality based on existing civil registration and vital statistics (CRVS) systems would be difficult for many countries. Data were sometimes not analyzed or produced at the desired frequency and most deaths were not registered in a timely way, risking loss to follow-up and registration.

In response, from 2020 through 2021, the Bloomberg Philanthropies Data for Health Initiative and Resolve to Save Lives (“the initiatives”) supported 13 countries in Latin America, Africa, and Asia with differing levels of CRVS digitalization and completeness to implement rapid mortality surveillance (RMS) (See Table 1 for details). This support included technical assistance to improve how countries rapidly record, retrieve, and analyze total mortality data using existing CRVS systems or other data sources when needed [1, 3]. Assistance included a) guidance on measuring total mortality by age, sex, place of usual residence, and place of death,b) recommendations for governments to build on existing surveillance and CRVS systems, and c) a tool for calculating excess deaths associated with COVID-19 [4]. These components enabled standardized, streamlined mortality reporting to be shared daily, weekly, or monthly with authorities such as emergency operations centers.

**Table 1.** Description of Rapid Mortality Surveillance by Country.

| Country | Community Component | Coverage | Level of Digitalization | Reporting Cadre | Frequency | Number of deaths recorded through 2022 |
| --- | --- | --- | --- | --- | --- | --- |
| 1. Bangladesh | Yes | National CRVS system and countrywide sample of 500 hospitals | Medium | Registrars and health assistants at the subdistrict level<br>Statisticians at each government hospital | Monthly with 3-month lag<br>Monthly totals reported with a 2- to 3-week lag | 658,069 |
| 2. Brazil | Yes | National CRVS system and Ministry of Health vital statistics systems | High | Health officials—mortality system technical team | 2-week lag | 2,685,388 -- |
|  | Yes | Sao Paulo state | High | Health officials—director of strategic information with support of University of Sao Paulo | 2-week lag |  |
| 3. Burkina Faso | No | 4 of 13 administrative regions of the country: Centre (Ouagadougou), Haut Bassin (Bobo Dioulasso), North Center, and Sahel | Low | Trained data collection officers | Weekly reporting—lag not measured | 9,452 |
| 4. Colombia | Yes | National cause of death investigation system | High | Physicians | Weekly reporting—lag not measured | 531,565 |
| 5. Ethiopia | Yes | 23 of 1,085 woredas | Medium | Health extension workers | Weekly—lag not measured | 100,698 |
| 6. Ghana | No | Sample of health facilities across three geographic zones | Low | Community-based surveillance volunteers | Weekly—lag not measured | 13,755 |
| 7. Liberia | No | National sample of health facilities | Low | Hospital staff | Weekly—lag not measured | 1,970 |
| 8. Malawi | Yes | Sample of health facilities and catchment areas across three cities | Low | Health facility staff and community health workers | Weekly—lag not measured | 29,266 |
| 9. Peru | No | National: SINADEF electronic medical certification of cause of death system capturing close to all death (completeness of above 70%)<br>Epidemiology Unit: confirmed COVID-19 cases | High | Physicians, data managers | Weekly | 354,753 |
| 10. Rwanda | No | National (all hospitals n=46 accounting for 30% of deaths pre-COVID) | High | Physicians, and hospital data managers | Real-time data entry; 4 to 6-week lag | 13,166 |
|  | Yes | All deaths reported through digitized CRVS system | High | Civil registrars | Weekly |  |
| 11. Senegal | No | National (all public hospitals n=39; completeness unknown) | Low | Physicians, data managers | Weekly | 29,696 |
| 12. Sierra Leone | Yes | National sample (25% of districts n=16) | Low | Facility health recorders; child health and mortality prevention surveillance staff; maternal mortality surveillance staff; community death registrars | Weekly | 8,744 |
| 13. Togo | No | National (6 health regions) | Low | Health records staff | Weekly | 12,364 |

This internal process evaluation of RMS implementation sought to identify:

- Major issues of implementation and how challenges were addressed.
- Barriers and facilitators to establishing a sustainable RMS system.
- Degree of success in producing timely and useful data.

We are not aware of another multi-country process evaluation of establishing mortality surveillance systems under pandemic conditions, particularly in settings of varying CRVS system completeness and digitalization. While some countries were more successful than others in generating and using this data for pandemic response, our goal was to identify barriers and facilitators of success, with the hypothesis that when successfully established, RMS can provide timely and useful counts of deaths, even in settings that lack strong CRVS systems and with the competing demands of the pandemic.

## Methods

Our qualitative process evaluation used semi-structured interviews with key people to learn about their RMS implementation experiences in 13 countries: Bangladesh, Brazil, Burkina Faso, Colombia, Ethiopia, Ghana, Liberia, Malawi, Peru, Rwanda, Senegal, Sierra Leone, and Togo. We sought participation from government focal points in all supported countries. This resulted in 16 participants:

- 13 men and three women.
- 11 government officials, primarily representing CRVS and statistics agencies, who were involved in the day-to-day process of setting up and implementing RMS.
- Five Vital Strategies in-country coordinators or consultants seconded to government departments for RMS work.

Interviews were conducted August–October 2021 with: two people in each of three countries (n = 6) and one person in each of the other 10 countries (n = 10). For each country, clearances and approvals were obtained from appropriate national research ethics bodies. No remuneration or inducement was offered, and participants were informed they could stop the interviews at any time.

Our evaluation encompassed data reporting, capture, and use, focusing on the RMS experience in countries where CRVS systems were not routinely capable of generating the data with the desired completeness and timeliness. Double-blind coding and qualitative analysis of interview transcripts using Dedoose (v 9.0.46) [5] surfaced themes that were categorized into key barriers and facilitators of RMS implementation.

It was not necessary or appropriate to have patients or the public involved in this research. To evaluate RMS implementation and achieve the desired goals of the research, it was necessary only to engage the staff who worked on this as part of their day-to-day responsibilities with their respective governments or public health agencies.

## Results

Five themes surfaced:

### Government ownership/buy-in

These were essential for successful implementation. We defined ownership as demand for and use of excess mortality data by those involved in pandemic response.

> At the beginning of the process, [the initiatives] gave us the idea…of asking [our country’s government] if they were interested in implementing a system of rapid surveillance of mortality. When we communicated it to the Ministry [of Health], we realized that it corresponded with a need that existed but wasn’t financed.
>
> —Hospital Director (Africa)^3^

We found that experience with mortality data analysis and awareness of its benefits motivated officials in Brazil, Colombia, and Peru to prioritize RMS. These countries were also more likely to have integrated existing mortality surveillance systems with CRVS systems and institutionalized processes for all-cause mortality analysis. Participants in all 13 countries were committed to RMS as part of the government’s duty to monitor public health.

> Mortality data reported on a weekly basis in real time would help us to plan and understand disease patterns, because if people are dying…we want to go out and investigate why. Then we’re able to stop that epidemic from escalating into a nationwide epidemic or global pandemic…That’s the reason the Director [of the National Health Service] was very much interested in the RMS strategy.
>
> —Technical Coordinator (Africa)

Making staff and resources available and assigning clear roles and responsibilities within ministries were key to successful implementation. Conversely, in countries with limited government buy-in, we observed implementation challenges. Lack of support by those involved with the pandemic response led to a) insufficient scope of the work and b) delay or discontinuation of the initiative because time and resources went to other pandemic-related responsibilities.

> Coordination of mortality surveillance is weak at all levels of the health system. There are no assigned persons responsible for mortality surveillance data reporting at the facility or community level. Ensuring that the purpose of institutionalizing mortality surveillance is understood by all relevant internal and external stakeholders is at the preliminary phase. —Government Surveillance (Africa)

### Data collection and digitalization

Completeness and digitalization of CRVS systems varied among the countries. Where systems were robust and digitalized, cause-of-death data could be seamlessly and rapidly incorporated into existing processes, so the CRVS system supported RMS. However, additional effort was often needed to capture data from remote areas and underserved populations.

> We still have some difficulties, especially in places where there is no easy access to the internet; [there,] they still have manual [cause-of-death] certification, but it is a very small percentage compared to the total number of cases that are presented… —Government IT Expert (Latin America)

Countries lacking robust CRVS systems encountered significant barriers to data collection, so parallel data collection mechanisms or procedures, such as purposive quota sampling of health facilities, had to be established (see Table 1). While these efforts were largely facility-based, they occasionally extended to capture deaths in the catchment areas.

> We didn’t have any structure or any functional [CRVS] system at the community level that would record deaths in the community and pass it on. What we have now are volunteers who are in the communities who pass on information on deaths that occur in their community. That was combined with deaths that would occur in the facility for that community. The facility…would then pass it on to the next level, and then the next level after that…and then…to [the capital]. — Medical Specialist (Africa)

### Interagency collaboration and data sharing

Effective collaboration among government agencies and other pandemic responders was essential for success. Where strong governance structures and clear data-sharing agreements existed, RMS data were more likely to be used effectively. Where such mechanisms were absent, collaboration and data sharing were often hindered. Establishing new relationships required deliberate efforts, with memoranda of understanding and standard operating procedures critical in facilitating cooperation. This underscored the importance of fostering collaboration, ensuring accountability, demonstrating leadership, and actively engaging with key pandemic responders to strengthen RMS systems.

In some countries, existing or lapsed inter-agency connections were strengthened or reestablished during the pandemic.

> The hospital epidemiologic surveillance network is an important structure that worked with death data but was suspended, [which] weakened this work. However, we are currently trying to re-establish this alliance with a better infrastructure because this is going to profoundly contribute to our work. These are spaces where we work a lot together since we cannot always be inside hospitals, they help us in this battle for information. — Government Surveillance (Latin America)

Other countries enhanced RMS by using other national health information systems to cross-check and validate mortality data. In one African country, linked databases for the CRVS systems and verbal autopsy facilitated validation of community deaths and generation of disaggregated mortality reports.

> The government has finished the integration of the civil registration system… which allows the entire birth/death registration, conducting verbal autopsies, and it provided the probable cause of deaths, which we enter into [the Ministry of Health’s health information system]. The integration of different systems has already been completed to link death notification, registration, certification, and to conduct the verbal autopsy. —Technical Partner (Africa)

In some countries, the lack of data-sharing agreements hindered these efforts.

> At some point, there were strides [there were attempts made to] link us with the [national statistics and demography agency]. Instead of taking more time getting data from data grids, we could get this data straight. All that had to be done was to put in place measures on how best we could share data, but unfortunately…those efforts did not materialize. —Government Statistician (Africa)

### Data analysis and interpretation

The participation of people with expertise in data analysis and RMS data triangulation was critical to data accuracy. One Latin American country’s capacity to triangulate data from multiple national and sub-national sources facilitated this validation process, highlighting the value of integrated data systems.

> There are comparisons of data on events of public health interest, not only COVID but also maternal mortality and perinatal mortality, with the databases of the surveillance system. Similarly, in terms of civil registration, there is also this cross-checking process [that involves] the Civil Registry, when it creates its databases; the Ministry of Public Health; and the National Administrative Department of Statistics, which…carries out the cross-checking…and communicates about missing information. — Government IT Expert (Latin America)

Understanding contextual factors—such as differences in age, sex, and location, and healthcare access—was essential for data analyses. This enabled identification of populations disproportionately affected by excess mortality. An innovation supporting these efforts was the Excel-based excess mortality calculator developed by the initiatives [3].

> I downloaded the data on a weekly basis, analyzed using Excel, and then entered on the excess mortality calculator shared with…the focal person and the designate for the [director of public health] and then we look at issues like, is the data talking to each other…are they corresponding…on a weekly basis for the first phase, from November [2020] to February [2021]. —Technical Coordinator (Africa)

### Data use

RMS data complemented other epidemiological measures, informing communication campaigns, government policies, and interventions such as lockdowns and vaccine distribution. Dissemination included updates to public dashboards and reports to national health ministries and emergency operations centers. Despite these successes, keeping RMS data representative and complete remained a significant challenge.

One Latin American country established a commission to identify COVID-19 deaths, enabling a more accurate assessment of the pandemic’s direct contribution to excess mortality.

> This information on the open data platform has allowed independent citizens to use the information and improve the health of different population groups. Another significant achievement is the use of mortality information for several publications linked to [deaths from external causes] or mortality between vaccinated and unvaccinated populations. — Technical Partner (Latin America)

In another Latin American country, the technical guidance package and excess mortality calculator were instrumental in developing an RMS database. The country back-calculated a historical baseline to compare against 2020 and 2021 and uploaded the data to a public dashboard, allowing residents and journalists to track excess mortality by week, disaggregated by municipality, age, and sex. These insights helped shape government decisions on COVID-19 safety protocols and underscored the importance of producing and using high-quality all-cause and excess mortality data.

> We are requested biweekly to present RMS data or results to the emergency operations committee…If after looking at the mortality trends, they [see] many people are dying, so [they say] can’t we put some…movement restriction measures, so that we can reduce the number of deaths per day? [I] am extremely happy that our results have been used in making some decisions regarding prevention measures. —Technical Partner (Africa)

Countries facing challenges in these areas reported delayed and/or suboptimal RMS outputs.

> The data we have isn’t complete at this time, because we’ve noticed with the rapid surveillance that the data at the community level has a lot of deaths that we haven’t taken into account…If we don’t pay attention, it’s possible that some people tell us that the data doesn’t reflect all of the mortality…that it only reflects the mortality at the hospital level…So, this gives us a great picture of hospital mortality, but it can’t give us a full picture of mortality at a general level. —Technical Partner (Africa)

The data also uncovered significant gaps in existing mortality surveillance through CRVS systems. In one African country, RMS data exposed a six-month reporting lag, so the Ministry of Health has sought to integrate mortality surveillance into the routine surveillance system. This example underscores how data can drive improvements in overall health reporting systems.

## Discussion

This evaluation highlighted several themes.

- **COVID-19’s Impact on Mortality Surveillance**: After WHO declared COVID-19 a pandemic, timely mortality data became essential for understanding its impact. In countries with limited diagnostic capacity, total and excess mortality were crucial metrics. Yet not all countries were able to produce timely, representative measures of total or excess mortality, especially those launching surveillance during the pandemic.
- **Government Buy-In**: Successful RMS implementation depended on strong government ownership and commitment to using mortality data to guide pandemic response.
- **Data Collection and Digitalization**: In countries with well-established CRVS systems, RMS was integrated smoothly, while countries with less experience faced data gaps and delays.
- **Interagency Collaboration**: RMS relied on strong collaboration among government agencies. Data-sharing agreements and cross-checking between systems, such as health and CRVS, improved data accuracy and utility.
- **Data Analysis and Use**: Skilled personnel were crucial for data analysis, especially in triangulating data from multiple sources. RMS data informed public health measures such as lockdowns and vaccination strategies.
- **Challenges and Improvements**: Despite the successes, challenges persisted, including data incompleteness, reporting delays, and lack of coordination. This highlighted the need for sustained investments in CRVS systems and better integration with pandemic response.

This process evaluation, to our knowledge, represents the first comprehensive assessment of rapid mortality surveillance implementation during a pandemic, across countries, each with varying levels of CRVS systems. Our findings underscore the critical role of government ownership, digital infrastructure, and interagency collaboration to establish RMS systems. Countries with more advanced digitalization and data-sharing were more successful, underscoring the importance of technology as a driver of success.

Countries with robust, well-integrated CRVS systems were better equipped to use RMS to produce timely mortality data. Strong integration between digitalized CRVS systems and national health surveillance frameworks was notably linked to supportive government leadership, clear roles, and defined responsibilities. And governments used RMS data to inform public health decision-making, revealing these systems’ value, particularly in pandemic response.

We also uncovered significant gaps in national systems, particularly in CRVS frameworks, for recording incident deaths. Barriers to effective RMS implementation included delays in data collection and issues with data completeness, compounded by data-sharing challenges, especially in countries with less-developed CRVS systems. These findings highlight the urgent need for sustained investment in CRVS systems and emphasize the importance of establishing robust governance structures to support RMS.

## Strengths and limitations

This evaluation has shown that countries appreciate the value of regular, routine analysis of death data to identify excess mortality early. Lockdowns in some countries disrupted vital statistics registration, rendering the collection of community-based notification and registration data impossible. Countries are keen to continue building and strengthening their mortality surveillance systems, integrating them into broader CRVS frameworks. Future government policies, public health strategies, and research should actively support these efforts.

This assessment also had some limitations. First, it may suffer from self-selection and self-reporting biases. Those who chose to participate in the interviews might have had strong views on RMS—either positive or negative—not fully representative of the governments’ perspectives. In terms of self-reporting, social desirability bias may have been at play. Second, our purposive sampling yielded a strong gender imbalance in the sample with 13 men and only 3 women. Lastly, interviews were conducted more than a year after the initial wave of COVID-19 during which many of the initial country RMS efforts were launched.

## Conclusion

The evidence supporting our hypothesis that RMS can provide effective mortality counts in low- and middle-income countries, even those with weak CRVS systems, was mixed. There was a broad appreciation of the value of mortality surveillance, and countries with fairly complete and digitalized CRVS systems were able to provide RMS data to government and to the public. However, many countries, particularly in Africa, struggled to produce facility data in a timely, complete, representative or scaled manner.

Given this mixed picture, the experience of implementing RMS during the COVID-19 pandemic offers crucial lessons for future public health surveillance efforts. A well-established, government-owned and -led system is essential for strengthening long-term capacity, spotlighting health disparities, making evidence-based decisions on resource allocation, and predicting future needs based on current data, historical trends, and patterns. We hope that these insights will guide the development of more resilient and responsive systems to produce measures of total and excess mortality.

## Data Availability

The interview transcripts underlying the results presented in the study are available from Vital Strategies, but were not necessary to be made public.

## Ethics Approval

This research followed the Vital Strategies protocol for Human Subjects Research (HSR), including submission of the research concept and interview guide for HSR status determination. The research was determined to be non-HSR based on the following “The proposed evaluation is not human subjects research because all information collected from respondents relates to the rapid mortality surveillance system in their country. Therefore, the evaluation does not involve “human subjects”.” As a result, there was no review or approval required from an Ethics Committee.

## Acknowledgements

Support for this project was provided by Vital Strategies’ Civil Registration and Vital Statistics and Data Impact programs through the Bloomberg Philanthropies Data for Health Initiative and Resolve to Save Lives. The authors thank the interviewees in the 13 countries for their time and insights; Victor Hugo Alvarez Castaño, Benjamin Clapham, Valens Nndagijimana Kagisha, Fatima Marinho, Raven Meade, Emmanuel Ngwakongnwi, Elizabeth Ortiz, and Moyeen Uddin, for assistance in study design and data collection and interpretation; Ellen Kuwana for assistance in drafting the manuscript; and Karen Schmidt for editorial support.

## Disclaimer

This work is an output of the Bloomberg Philanthropies Data for Health Initiative (www.Bloomberg.org) and Resolve to Save Lives. The views expressed in the submitted article are the authors’ own and not an official position of the Philanthropies.

## Footnotes

3 To preserve the anonymity of sources, we refer only to the region of the respondent.

